# Empowering Research on Epilepsy Surgery Outcomes

**DOI:** 10.1101/2022.05.11.22274965

**Authors:** Adam S. Dickey, Robert T. Krafty, Nigel P. Pedersen

## Abstract

Low statistical power is a recognized problem in many fields. We performed a systematic review to determine the median statistical power of studies of epilepsy surgery outcomes. We performed a PubMed search for studies reporting epilepsy surgery outcomes for the years 1980-2020, focusing on studies using stereoelectroencephalography (SEEG). We extracted patient count data for comparisons of surgical outcome between two groups, based on a reported prognostic factor. We defined a clinically meaningful difference as the difference in seizure freedom for MRI positive (66.9%) versus negative (45.5%) from the largest study found. Based on 69 studies of surgery outcomes in patients undergoing SEEG, the median sample size was 38 patients, and the median statistical power was 24%. This implies at least a 17% chance a study with a significant result is false, assuming 1:1 pre-test odds. Results from simulation studies suggest that, if a typical SEEG study finds a significant effect, then the median observed effect size will be more than double the true effect size. We conclude that studies of epilepsy surgery outcomes using SEEG are often statistically underpowered, which limits the reproducibility and reliability of the literature. We discuss how statistical power could be improved.

**SHORT SUMMARY:** We performed a systematic review to determine the median statistical power of studies of epilepsy surgery outcomes, focused on stereoelectroencephalography. We extracted patient count data for comparisons of outcomes between two groups. We defined a clinically meaningful difference as the prognostic value of a normal versus abnormal MRI. Based on 69 studies, the median sample size was 38 patients, and the median statistical power was 24%. Underpowered studies will overestimate the size of true effects and are more likely to report false positive results. We discuss how statistical power, and thus reproducibility and reliability of results, can be improved.

## 1 INTRODUCTION

Low statistical power is a recognized problem in many fields^1^, which means true effects will be missed, the chance a study with a significant result is false will be inflated^2^, and discovered true effects will be over-estimated^3^. In neuroscience the typical power to detect a true effect was estimated at 20%^4^. We recently highlighted the issue of low power in a single study of laser surgery^5^, but we suspected that low power was pervasive in studies of epilepsy surgery outcomes. We performed a systematic review to acquire a representative sample of papers reporting epilepsy surgery outcomes and determine the power of a typical study. We focus on stereoelectroencephalography (SEEG) to keep the review tractable, and because the recent adoption of this technique in the United States may lead to smaller number of patients and thus lower power.

A major challenge in any power analysis is determine the “clinical meaningful” effect size. This can be defined as how large of an effect we expect actually exists and that we want to be able to detect^1.^. The largest study in the SEEG literature we know of is from the Milan group, which reported outcomes of freedom from disabling seizures following surgery in 470 patients^6^. We use this large cohort to estimate a clinical meaningful effect size as the (statistically significant) prognostic value of an abnormal MRI: 66.9% (204/305) of patients with an abnormal MRI were seizure-free, compared to 45.5% (75/165) patients with a normal MRI (odds ratio 2.4, 95% CI 1.6-3.5). The presence of a lesion on MRI is not the only relevant prognostic factor but is commonly reported in the literature.

In our meta-analysis, we extracted the number of patients who were divided into two groups (such as MRI positive or negative) and computed the power of that study to predict a difference in seizure freedom of 66.9% versus 45.5%. We review options for computing power. We discuss the issues raised by underpowered studies and highlight options for improving power.

## 2 METHODS

### 2.1 Literature Search

We performed a PubMed search with the terms “((SEEG) OR (stereoelectroencephalography)) AND (epilepsy) AND (surgery outcome)” for the year of publication from 1980 to 2020, last performed March 2, 2021. One of us (ASD) reviewed each abstract, and then examined the full manuscript when the abstract described a comparison of surgery outcome between two groups. Inclusion criteria were 1) studies of patients undergoing SEEG that 2) report a comparison of surgical outcome (good or bad) based on a prognostic factor. Exclusion criteria included reviews, small case series, or mixed populations of SEEG and subdural grids. Raw patient counts were extracted for the comparison most emphasized in the abstract. If raw patient counts were not reported as a two-by-two table, we inferred the patient counts from information given in the text. Patient counts for each included study are reported in the supplement (Appendix S1).

### 2.2 Calculation of power

We estimated the power of the Chi-square test for a difference in proportions using three methods. First, we simulated binomial count data for each group using our predefined plausible effect size^6^ (66.9% vs. 44.5%). We simulated 10,000 repetitions and report the power as the percentage of repetitions where a Chi-square test reported a p-value ≤ 0.05. We did this for each study using the reported number of patients in each group. This software analysis was done in MATLAB (version R2016b, Mathworks, Natick, MA) using the function **chisquarecont**, available on the MATLAB Central File Exchange^7^.

Second, we estimated the statistical power of the Chi-square test by approximation^8^ (Equation 1) using the Gaussian cumulative density function (*Φ*) applied to a critical value, which is a function of the sample size (*n1* and *n2*) and percent seizure free (*p1* and *p2*)of each group, and the Z-score *z*_*α/2*_ (1.96 if *α* = 0.05).

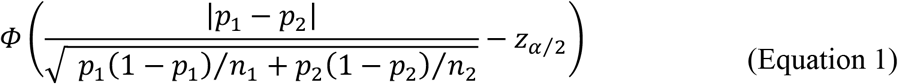

Finally, we used the R package “Exact” and the function **power.exact.test** using method “pearson chisq” (R version 4.1.1, www.r-project.org) to compute power. Traditionally, Fisher’s exact test is recommended if there is an expected cell count is less than 5, but is this is known to have lower power relative to a Chi-square test (and the equivalent Z-test) for many trial models^9^. For simplicity, we only consider Chi-square test, since if anything this will overestimate power relative to a mixture of Chi-square and Fisher’s exact tests.

### 2.3 Chance a study with a significant result is false

A p-value is the probability of obtaining a test statistic from an independent study as or more extreme than the statistic from the observed study, assuming the null hypothesis is true^10^. That is, the p-value is a probability based on observed data given the null hypothesis. The p-value does not measure the probability that the data were produced by chance alone, or the probability the null hypothesis is true^11^, which is a probability of the null hypothesis given the data. Bayes’ Rule can be used to convert the probability of data given the null into the probability of the null given data. This requires the pre-study odds that the null hypothesis is true and, crucially, the statistical power. The positive predictive value (PPV) is given by the equation below, where (1-*Beta*) is the power, *Beta* is the Type II error, *Alpha* is the Type I error, and *R* is the pre-test study odds^4^ (Equation 2)

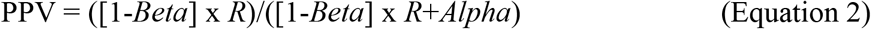

We assume the pre-test odds *R* are 1:1, meaning the pre-test probability of the alternative hypothesis being true is 50%. This is the most optimistic plausible scenario. If the pre-test probability of the alternative is less than 50%, the PPV will be even loweR^2^. The chance a study with a significant result is false is one minus the PPV (Equation 3).

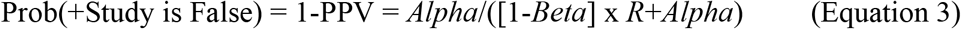

### 2.4 Over-estimation of effect size

It is known that if a study is underpowered, then the an observed effect size will on average overestimate the true effect size if it is found to be statistically significant^3^. We define a “typical” SEEG study has having the median sample size with the median allocation ration across all included studies. Using the simulation paradigm described above, we quantify the over-estimation of a typical study by computing the median odds-ratio of runs with a Chi-square p-value <0.05, divided by the assumed odds ratio of 2.4.

## 3 RESULTS

We reviewed 119 manuscripts in detail, from the 268 abstracts returned by our search terms (and one added from citations). 69 studies met inclusion criteria, meaning they reported a comparison of good versus poor surgical outcome based on some prognostic factor in patients undergoing SEEG.

The median sample size was 38 patients (range 8-470). We estimated power using our assumed effect size of 66.9% vs. 45.5% seizure freedom between two groups. The median power using a chi-square test was 24% (range 7-99%) using the exact calculation using R or the simulation-based approach. The power was 22% using the Gaussian approximation, which slightly under-estimated the true power (Figure 1A). A power of 24% corresponds to a 17% (0.05/[0.24+0.05]) chance that a study with a significant result (at p<0.05) is false (Figure 1B).

**Figure 1:**
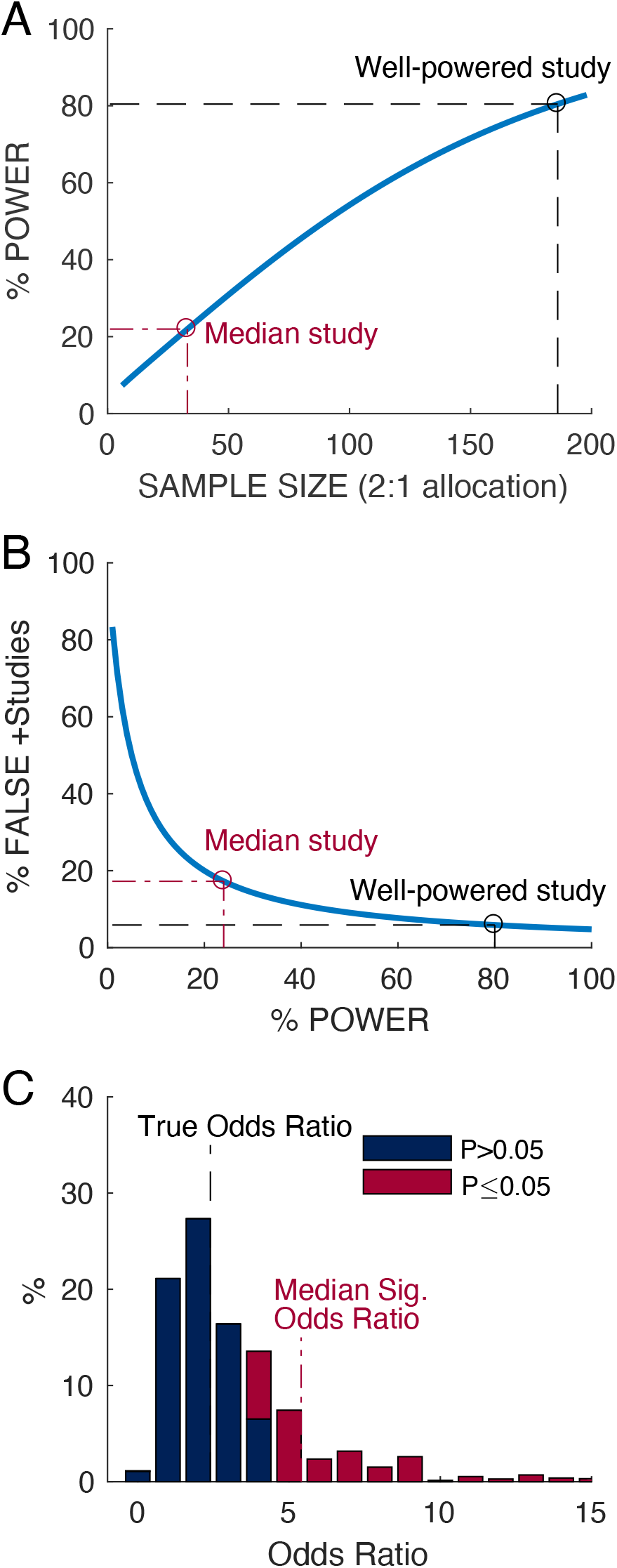
**A) Statistical power compared to sample size** Assuming 2:1 allocation, a study would need 186 subjects (124 vs. 62) to have 80% statistical power to detect a difference in seizure freedom of 66.9% in group 1 and 45.5% in group 2 (odds ratio 2.4). However, the median SEEG study has only around 24% power to detect this difference. **B) Statistical power compared to chance study is false**. Assuming a pre-test probability of 50% that the alternative hypothesis is true and a statistical test with alpha = 0.05, the chance a study with 80% statistical power which a significant result is false is 5.9%. However, the median EEG study with 24% statistical power with a significant result has a 17% chance of being false. **C) Median odds ratio for significant versus non-significant results**. We defined a “typical” SEEG study as having the median sample size (38) with the median allocation percentage (63%), which rounded to 24 subjects in group 1 and 14 in group 2. When this experiment is simulated, the median odds ratio that is significant at p<0.05 is 5.4, which is more than double the true odds ratio of 2.4. Thus a typical underpowered study, if it finds an significant result, will overestimate the effect size.

The median allocation percentage was 63% of the patients in the positive group (roughly a 2:1 ratio). We therefore define a “typical” SEEG study as having median sample size (38) with the median allocation percentage (63%), which rounds to 24 patients in the positive group and 14 in the negative group. Based on simulations, the median odds ratio that was significant at p<0.05 was 5.4, which is more than double the assumed true odds ratio of 2.4 (Figure 1C). Thus, when a typical SEEG study finds a significant effect, its magnitude will be over-estimated.

Though clinical trials are often designed to achieve 80% statistical power, only 3% (2/69) of the surveyed studies had greater than 80% power to detect our assumed effect size. We highlight the seven studies which had more than 50% power to detect our assumed effect size (Table 1).

**Table 1:** Selected SEEG studies with greater than 50% statistical power. Power was calculated assuming to detect a difference in seizure freedom of 66.9% in group 1 and 45.5% in group 2 (Odds Ratio 2.4). Only 7/69 (10%) of studies have more than 50% power. Column titles report the first author (Author), year of publication (Year), total sample size (Ntot) and size of the positive (N1) and negative group (N2), the estimated power (Power), the finding present in the positive group (+ Finding), surgical outcome (Outcome), and the percentage of patients with seizure reduction in the positive (SR+) or negative group (SR-). Abbreviations: MRI magnetic resonance imaging, LFS low frequency stimulation, ECOG electrocorticography, RF-TC radiofrequency thermocoagulation.

| Author | Year | Ntot | N1 | N2 | Power | (+) Finding | Outcome | SR (+) | SR (-) |
| --- | --- | --- | --- | --- | --- | --- | --- | --- | --- |
| Cardinale<sup>6</sup> | 2019 | 470 | 305 | 165 | 99% | MRI lesion | Engel I | 67% | 45% |
| Trebuchon<sup>12</sup> | 2020 | 346 | 120 | 226 | 97% | Seizure induced with LFS | Engel I | 72% | 59% |
| Cossu<sup>14</sup> | 2005 | 165 | 123 | 42 | 70% | MRI lesion | Engel I | 63% | 38% |
| Grewal<sup>15</sup> | 2019 | 127 | 75 | 46 | 65% | Full ECOG field resection | Excellent | 65% | 52% |
| Lagarde<sup>16</sup> | 2018 | 143 | 114 | 29 | 57% | Low-voltage fast activity | Engel I | 60% | 31% |
| Bourdillon<sup>17</sup> | 2017 | 91 | 44 | 47 | 56% | Favorable initial RF-TC | Good | 93% | 55% |
| Tassi<sup>18</sup> | 2012 | 100 | 66 | 34 | 55% | Balloon cell histology | Engel I | 88% | 74% |

## 4 DISCUSSION

Based on a representative sample of 69 studies of epilepsy surgery outcomes in SEEG, we estimate that the median study has only 24% power to detect a clinically meaningful effect. Though we focused on SEEG, we expect low power is common in studies of epilepsy surgery. An underpowered study (by definition) has a low chance to detect a true effect when present. However, some studies incorrectly interpret absence of evidence as evidence of absence. If an underpowered study finds no statistical difference in seizure freedom between an MRI positive and MRI negative group, then it is incorrect to conclude that MRI could not at least partially predict seizure freedom. If the study power is less than 50%, a negative result is expected even if the effect is real.

Another statistical fallacy is treating a result significant at p<0.05 as only having a 5% chance of being false^11^. The median SEEG study with 24% power has at least a 17% chance of being false, based on power considerations alone. Moreover, if an underpowered study finds a significant effect, it will over-estimate the effect size. Such limitations of low power are rarely discussed in epilepsy literature.

The biggest limitation of this study is our reliance on a single study of MRI positive vs. negative to determine the clinically meaningful effect size. There are countless possible prognostic factors, which may have different true effect sizes, which will be need quantified in future (ideally well-powered) studies. Our recommendation is that authors perform their own formal statistical power analysis, with an explicit definition of the clinically meaningful effect size. If the study is underpowered, that should be discussed in the interpretation of the results.

The most obvious (and most difficult) way to increase power is to include more subjects. This can be done by combining data across institutions, as done by one study highlighted here^12^. A less obvious way to increase power is to expand statistical analysis beyond the comparison of two-by-two tables of binary predictors and surgical outcomes. For example, we have shown that using ordinal regression using the full ordinal Engel surgical outcomes increases power relative to traditional binary logistic regression^13^. A similar increase in power should be possible by using continuous or ordinal predictors, rather than binary predictors.

## 5 CONCLUSION

Studies of epilepsy surgery outcomes in SEEG are systematically underpowered. Future studies should perform formal power analyses, and low power should be addressed by increasing the sample size and/or by statistical analysis which avoids unnecessary dichotomization. We call for increased statistical rigor in studies of surgery outcomes, as well as greater collaboration within the epilepsy community, with the goal of providing accurate and precise information to guide treatment for our patients with epilepsy.

## Data Availability

MATLAB and R code which can be used to reproduce the analyses and figures described here will be posted at https://github.com/adamsdickey.

## FUNDING AND ACKNOWLEDGEMENTS

N.P.P. is supported by the Woodruff Foundation, CURE Epilepsy, and NIH grants K08 NS105929, R01 NS088748, and R21 NS122011. A.S.D. is supported by the National Center for Advancing Translational Sciences of the NIH under award number UL1 TR002378 and KL2 TR002381. R.T.K. is supported by R01 GM113243. The content is solely the responsibility of the authors and does not necessarily represent the official views of the National Institutes of Health. MATLAB and R code which can be used to reproduce the analyses and figures described here will be posted at https://github.com/adamsdickey.

## AUTHORSHIP STATEMENTS

A.S.D and N.P.P. contributed to the conception and design of the study. A.S.D, R.T.K and N.P.P contributed to the drafting of the text. A.S.D. performed the statistical analysis and prepared the figures and tables

## TABLE

**Appendix S1:**
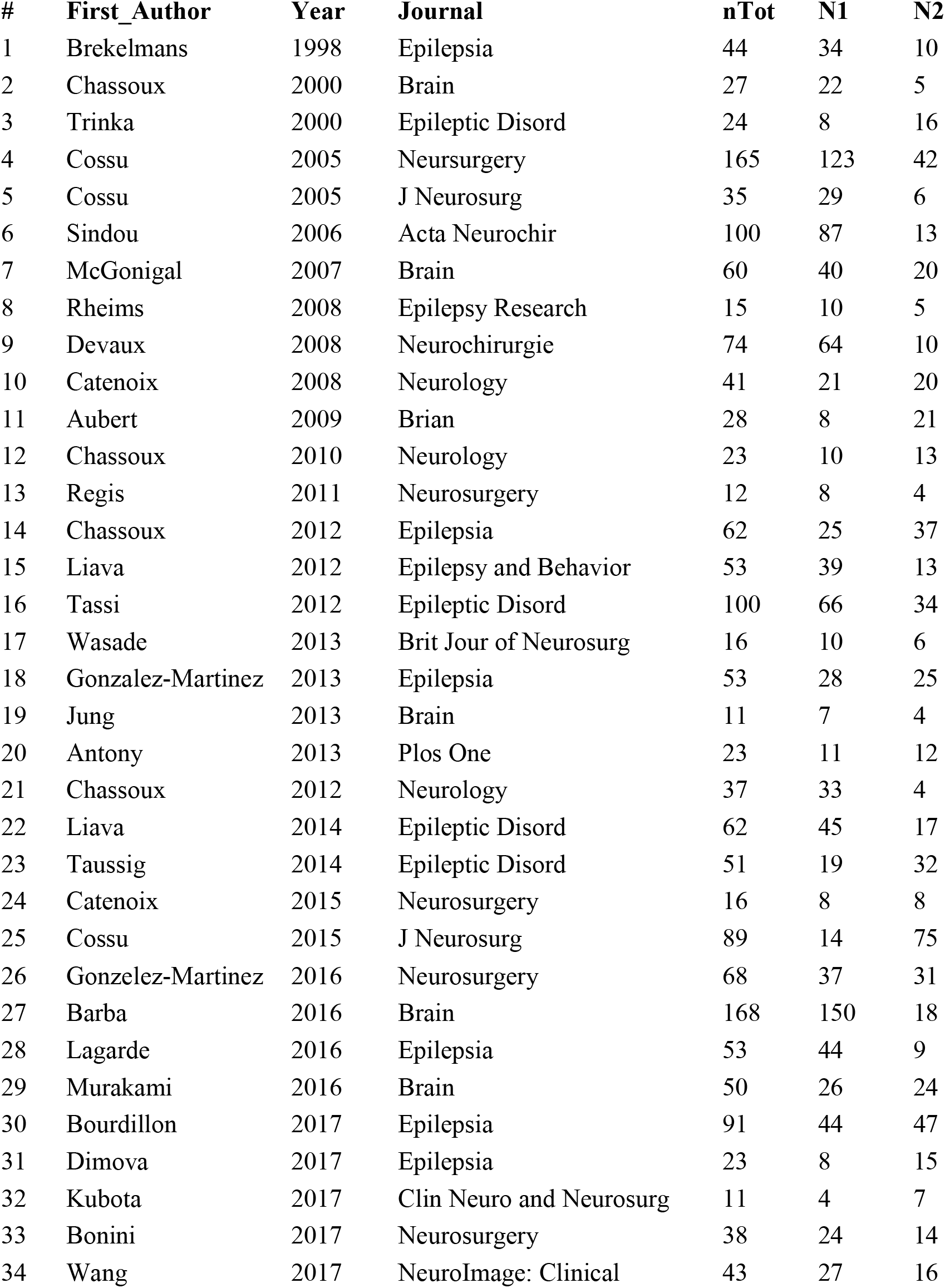
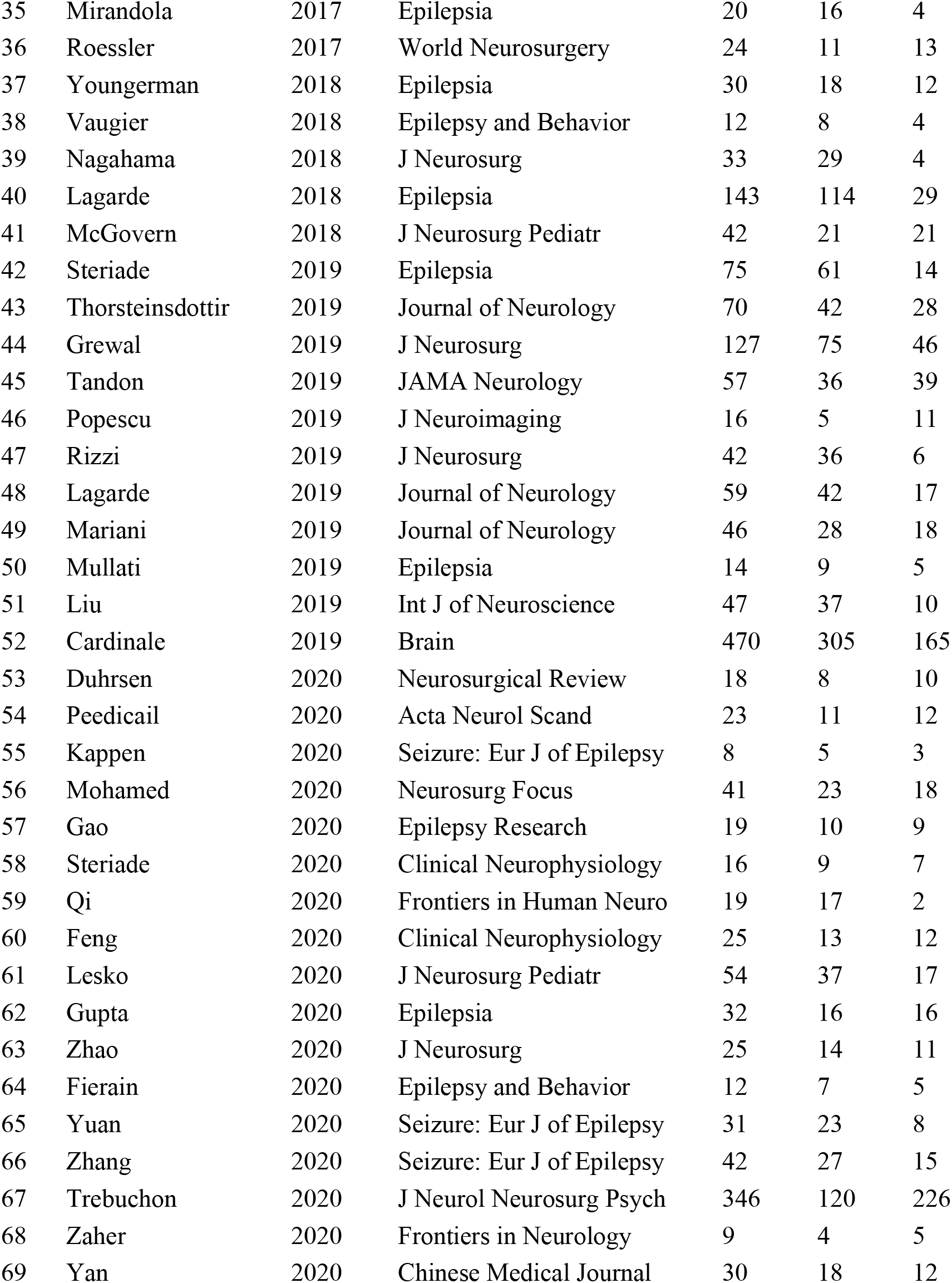
Raw patient counts from 69 studies. For left to right, columns list the paper ID (#), the first author (First Author), year of publication (Year), journal of publication, (Journal), the total sample size (nTot) and the size of the positive (N1) and negative (N2) prognostic groups.

